# Development and Validation of a Metabolite Index for Obstructive Sleep Apnea across Race/Ethnicities

**DOI:** 10.1101/2022.05.25.22275577

**Authors:** Ying Zhang, Debby Ngo, Bing Yu, Neomi A. Shah, Han Chen, Alberto R. Ramos, Phyllis C. Zee, Russell Tracy, Peter Durda, Robert Kaplan, Martha L. Daviglus, Stephen S. Rich, Jerome I. Rotter, Jianwen Cai, Clary Clish, Robert Gerszten, Bruce S. Kristal, Sina A. Gharib, Susan Redline, Tamar Sofer

**Affiliations:** Department of Medicine, Division of Sleep Medicine and Circadian Disorders, Brigham and Women’s Hospital, Boston, MA, 02115; Cardiovascular Institute, Division of Pulmonary, Critical Care and Sleep Medicine, Beth Israel Deaconess Medical Center, Boston, MA, 02215; Human Genetics Center, Department of Epidemiology, Human Genetics and Environmental Sciences, School of Public Health, The University of Texas Health Science Center at Houston, Houston, TX, 77030; Department of Medicine, Albert Einstein College of Medicine, New York, NY, 10029; Center for Precision Health, School of Biomedical Informatics, The University of Texas Health Science Center at Houston, Houston, TX, 77030; Department of Neurology, Sleep Medicine Program, University of Miami Miller School of Medicine, Miami, FL, 33136; Department of Neurology, Division of Sleep Medicine, Northwestern University, Chicago, IL, 60611; Department of Pathology Laboratory Medicine, Larner College of Medicine, University of Vermont, Burlington, VT, 05405; Department of Epidemiology & Population Health, Albert Einstein College of Medicine, Bronx, NY, 10461; Department of Preventive Medicine, Northwestern University Feinberg School of Medicine, Chicago, IL, 60612; Center for Public Health Genomics, University of Virginia, Charlottesville, VA, 22908; Department of Pediatrics, The Institute for Translational Genomics and Population Sciences, The Lundquist Institute for Biomedical Innovation at Harbor-UCLA Medical Center, Torrance, CA, 90502; Department of Biostatistics, Collaborative Studies Coordinating Center, University of North Carolina at Chapel Hill, Chapel Hill, NC, 27599; Metabolite Profiling Platform, Broad Institute of MIT and Harvard, Cambridge, MA, 02142; Cardiovascular Research Center, Massachusetts General Hospital, Harvard Medical School, Boston, MA, 02115; Department of Medicine, Sleep and Circadian Disorders, Harvard Medical School, Boston, MA, 02115; Department of Medicine, Division of Sleep Medicine and Circadian Disorders, Brigham and Women’s Hospital and Harvard Medical School, Boston, MA, 02115; Department of Medicine, Division of Pulmonary, Critical Care, and Sleep Medicine, University of Washington, Seattle, WA, 98195; Department of Biostatistics, Harvard T.H. Chan School of Public Health, Boston, MA, 02115

## Abstract

**Background:** Obstructive sleep apnea (OSA) is a common disorder characterized by recurrent episodes of upper airway obstruction during sleep resulting in oxygen desaturation and sleep fragmentation, and associated with increased risk of adverse health outcomes. Metabolites are being increasingly used for biomarker discovery and evaluation of disease processes and progression. Studying metabolomic associations with OSA in a diverse community-based cohort may provide insights into the pathophysiology of OSA. We aimed to develop and replicate a metabolite index for OSA and identify individual metabolites associated with OSA.

**Methods and Findings:** We studied 219 metabolites and their associations with the apnea hypopnea index (AHI) and with moderate-severe OSA (AHI≥15) in the Hispanic Community Health Study/Study of Latinos (HCHS/SOL) (n=3507) using two methods: (1) association analysis of individual metabolites, and (2) least absolute shrinkage and selection operator (LASSO) regression to identify a subset of metabolites jointly associated with OSA, and develop a metabolite index for OSA. Results were validated in the Multi-Ethnic Study of Atherosclerosis (MESA) (n=475). When assessing the associations with individual metabolites, we identified seven metabolites significantly positively associated with OSA in HCHS/SOL (FDR p<0.05), of which four associations - glutamate, oleoyl-linoleoyl-glycerol (18:1/18:2), linoleoyl-linoleoyl- glycerol (18:2/18:2) and phenylalanine, replicated in MESA (one sided-*p* <0.05). The OSA metabolite index, composed of 14 metabolites, was associated with 50% increase of risk for moderate-severe OSA (OR=1.50 [95% CI: 1.21-1.85] per 1 SD of OSA metabolite index, *p*<.001) in HCHS/SOL and 44% increased risk (OR=1.55 [95% CI: 1.10-2.20] per 1 SD of OSA metabolite index, *p*=0.013) in MESA, both adjusted for demographics, lifestyle, and comorbidities. Similar albeit less significant associations were observed for AHI.

**Conclusions:** We developed a metabolite index that replicated in an independent multi-ethnic dataset, demonstrating the robustness of metabolomic-based OSA index to population heterogeneity. Replicated metabolite associations may provide insights into OSA-related molecular and metabolic mechanisms.

## Introduction

Obstructive sleep apnea (OSA) is a common disorder characterized by recurrent episodes of upper airway obstruction during sleep resulting in oxygen desaturation and sleep fragmentation (1). While highly prevalent in the population (2), OSA is severely under- diagnosed (3), especially in women (4). For example, only 1.3% of participants in the Hispanic Community Health Study/Study of Latinos (HCHS/SOL) and 7-15% of participants in the Multi-Ethnic Study of Atherosclerosis (MESA) reported a previous OSA diagnosis; in MESA, underdiagnosis was highest among race/ethnic minorities (5,6). The pathophysiology underlying OSA is multifaceted, which includes obesity, craniofacial structure, upper airway neuronal control, ventilatory control, and inflammation, among others (7). OSA is also associated with increased risk of adverse health outcomes, including hypertension, cardiovascular disease, diabetes, and early mortality (8-11).

Metabolomics is the study of small biochemical compounds at large scales (12). As a growing number of large metabolomics datasets become available, metabolites are being increasingly leveraged for biomarker discovery and evaluation of disease processes and progression (13,14). In particular, studying metabolite associations with OSA may improve our understanding of the pathophysiology of OSA. However, research in this area has been limited by the relatively small number of participants that have undergone both overnight sleep studies and metabolomic profiling, the lack of representativeness of the study subjects selected solely in clinical encounters, and the limited metabolite panels used by many targeted metabolomics platforms (15,16).

Here, we study metabolite associations with OSA in the HCHS/SOL, one of the largest multi- center cohorts with diverse participants from a rapidly growing minority group in the US: Hispanics/Latinos. We then test these associations for replication in MESA, a multi-ethnic community-based cohort. Our study follows the design described in **Figure 1**. First, we study associations of individual metabolites with a measure of moderate to severe OSA, defined by the apnea hypopnea index AHI ≥ 15, as well as by associations with continuously measured AHI. Next, we develop a metabolite index for OSA by aggregating together multiple metabolites as a potential biomarker for OSA. We then validate its association with OSA in MESA. This process is repeated for a continuous measure of OSA severity, the AHI. Because OSA has different characteristics across sexes (17), in secondary analyses we additionally studied sex-specific metabolite indices.

**Figure 1:**
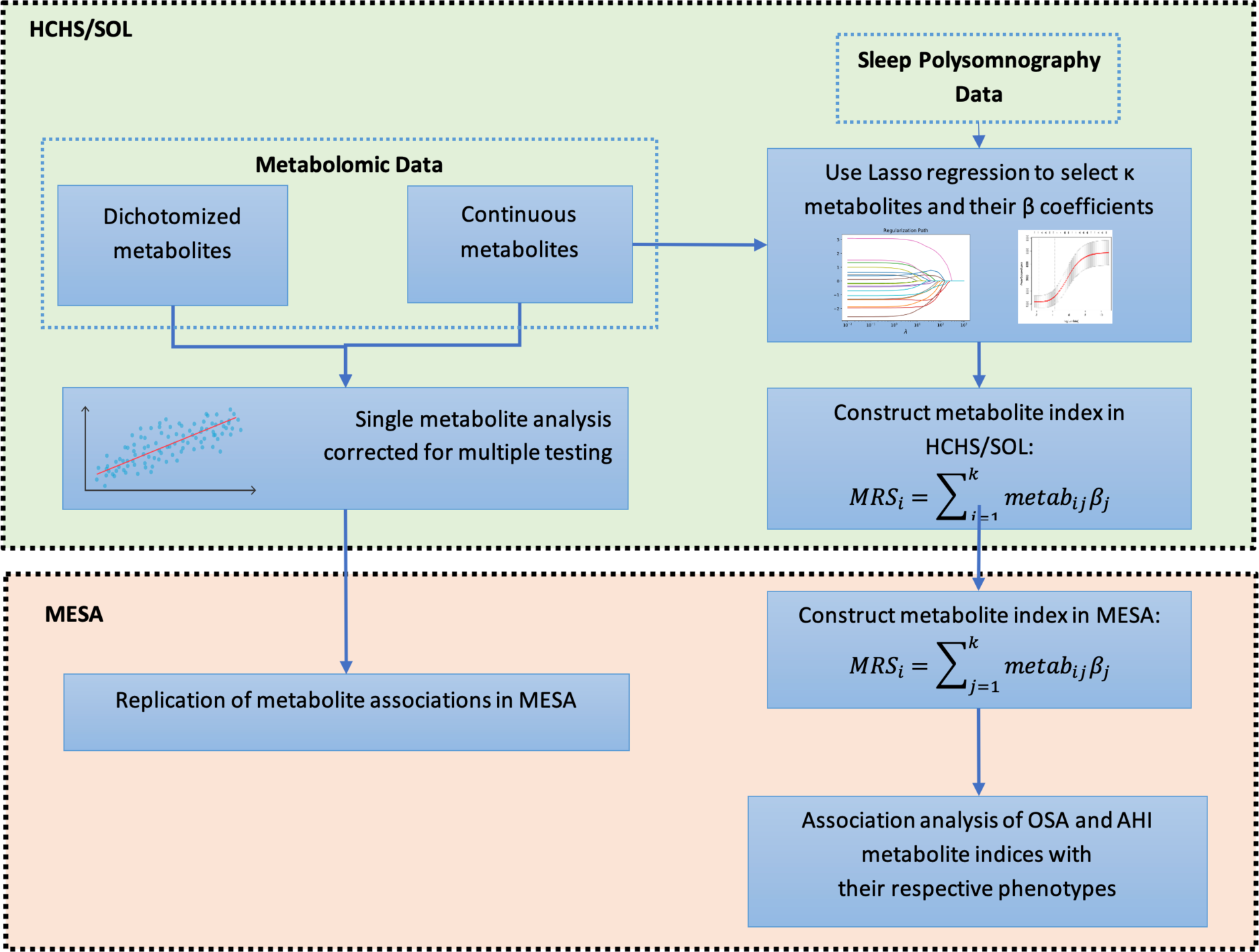
Study design flow chart. Definition of abbreviations: LASSO = least absolute shrinkage and selection operator, HCHS/SOL= the Hispanic Community Health Study/Study of Latinos, MESA= the Multi-Ethnic Study of Atherosclerosis.

## Methods

### The Hispanic Community Health Study/Study of Latinos

The HCHS/SOL is a community-based cohort study of 16,415 self-identified Hispanic/Latino persons from diverse Hispanic/Latino backgrounds (Mexicans, Puerto Ricans, Cubans, Central Americans, Dominicans, and South Americans) (18). Participants 18–74 years of age at their baseline examination were recruited through a stratified multistage area probability sample design from four communities: San Diego, California; Chicago, Illinois; The Bronx, New York; and Miami, Florida. The baseline examinations occurred in June 2008–July 2011 and included assessment of OSA using a validated Type 3 home sleep apnea test (ARES Unicorder 5.2; B-Alert, Carlsbad, CA) that measured nasal air-flow, position, snoring, heart rate and oxyhemoglobin saturation (19) as previously described (5) in 14,440 individuals. The current cross-sectional analysis included 3,507 of these participants who also had blood assessed for metabolomic measures. Our primary analysis focused on a metabolomics-based biomarker for moderate or severe OSA defined as AHI≥15, with events defined as apneas or hypopneas with at least 50% cannula flow reduction for a minimum duration of 10 seconds, with ≥3% oxygen desaturation. The sleep study was conducted within the week following the baseline exam in which blood was collected and used for metabolite quantification. The study was approved by Institutional Review Boards at each field center where all participants provided written consent, and by the study’s Reading and Data Coordinating Centers.

### HCHS/SOL Metabolomic Profiling

Fasting blood samples were collected at the baseline examination. Of all HCHS/SOL participants from the baseline examination who also had genetic data, 3,968 individuals were selected at random for metabolomics assessment. The samples were processed, and serum was stored at −70°C since collection. The metabolomic profiling was conducted at Metabolon (Durham, NC) with Discovery HD4 platform in 2017. Serum metabolites were quantified with untargeted, liquid chromatography-mass spectrometry (LC-MS)-based quantification protocol (20). The platform captured a total of 1,136 metabolites, including 782 known and 354 unknown (unidentified) metabolites.

### The Multi-Ethnic Study of Atherosclerosis

MESA is a cohort study designed to study risk factors for clinical and subclinical cardiovascular diseases in four racial/ ethnic groups (21). The study began in July 2000 and recruited 6,814 adults free of clinical CVD and aged 45–84 years from 6 centers: Baltimore, MD; Chicago, IL; Los Angeles, CA; New York, NY; Saint Paul, MN; and Winston-Salem, NC. Participants were continued to be studied through subsequent follow-up exams. Of the 4,077 participants who attended Exam 5 (2010-2012), 2,261 participated in the MESA Sleep ancillary study (2010- 2013). As reported before (6), participants in the Sleep Exam were generally similar to non-participants. OSA was assessed with Type II in-home polysomnography. AHI was defined as the total number of apnea and hypopneas with at least 30% reduction in the nasal flow signal and with ≥ 3% oxygen desaturation per hour of sleep. The median time interval between Exam 5 fasting plasma sample collection and MESA Sleep was 301 days (range 0–1,024 days). Metabolomic data was collected on 1,000 randomly selected participants from Exam 5. Of these, 475 participants also had sleep measures and are included in this analysis. Local institutional review boards at all the participating institutions approved study protocols, and all participants gave written informed consent.

### MESA Metabolomic Profiling

Metabolite profiling was performed using liquid chromatography tandem mass spectrometry (LC-MS). Positive ion mode profiling of water-soluble metabolites and lipids was performed using LC-MS systems comprised of Nexera X2 U-HPLC (Shimadzu Corp.; Marlborough, MA) units coupled to a Q Exactive mass spectrometer (Thermo Fisher Scientific; Waltham, MA). Polar metabolites were analyzed using hydrophilic interaction liquid chromatography (HILIC) and lipids were analyzed separately using reversed phase C8 chromatography as described in detail previously (22). Raw data were processed using TraceFinder 3.1 (Thermo Fisher Scientific; Waltham, MA) and Progenesis QI (Nonlinear Dynamics; Newcastle upon Tyne, UK). To measure organic acids and other intermediary metabolites in negative ionization mode, chromatography was performed using an Agilent 1290 infinity LC system equipped with a Waters XBridge Amide column, coupled to an Agilent 6490 triple quadrupole mass spectrometer. Metabolite transitions were assayed using a dynamic multiple reaction monitoring system. LC-MS data were analyzed with Agilent Masshunter QQQ Quantitative analysis software. Isotope labeled internal standards were monitored in each sample to ensure proper MS sensitivity for quality control. Pooled plasma samples were interspersed at intervals of 10 participant samples for standardization of drift over time and between batches. Additionally, separate pooled plasma was interspersed at every 20 injections to determine coefficient of variation for each metabolite over the run. Peaks were manually reviewed in a blinded fashion to assess quality. For each method, metabolite identities were confirmed using authentic reference standards or reference samples. Metabolites with poor peak quality and coefficients of variation greater than 30% averaged across batches were removed from analysis.

### Quality control of metabolites in HCHS/SOL and MESA

Missing metabolite values were addressed as described in **Supplemental Figure 1**. In our discovery sample (HCHS/SOL), we excluded individuals with more than 25% missing metabolite levels, and metabolites with missing values for 75% or more individuals. For metabolites with more than 25% and less than 75% missing values, values were dichotomized as “observed” and “unobserved”. For metabolites with less than 25% missing values, we imputed the missing values using the minimum observed value of the metabolite in the sample, under the assumption that metabolites were not observed due to a technical detection limit.

Because our study design includes validation analysis, we focused on metabolites available in both HCHS/SOL and MESA. Before any quality control (QC) methods were applied, 231 HCHS/SOL metabolites were mapped to 294 metabolites in MESA. The mapping of MESA to HCHS/SOL metabolites as well as to RefMet ID was done at Clish Lab. MESA had multiple metabolites matched to a single HCHS/SOL metabolite in multiple instances because the same metabolite was measured via more than one platform used by MESA. In some cases, a single metabolite appears as two highly correlated ion features in the same MESA platform (e.g., some neutral lipids were measured as both sodium and ammonium adducts). Therefore, a single feature was mapped to the metabolite in HCHS/SOL while the redundant features were dropped according to the following principles: features with redundant ions were excluded; features with lower missingness and lower skewness were prioritized. After removing 60 such redundant features in MESA and applied QC methods based on the missingness in HCHS/SOL, 219 HCHS/SOL metabolites were mapped to 219 metabolites in MESA. **Table S1** provides the list of the 294 initially matched metabolites cross-referenced by RefMet ID and metabolite annotations including HMDB IDs provided by Metabolon, along with details regarding metabolite-specific QC resulting in the final list of one-to-one matched metabolites. The serum concentration values of the matched metabolites that were treated as continuous were rank-normalized.

Because MESA was a validation study, we only evaluated metabolites that were identified in the association analysis in HCHS/SOL. The missing data for these metabolites were always <25%, so we treated these as continuous variables and imputed missing values with the minimum observed value in the MESA sample.

### Statistical analysis

Association analyses were based on three conceptual regression models: Model 1 (i.e., primary model) adjusted for demographic variables – age, sex, study center, Hispanic background (Mexicans, Puerto Ricans, Cubans, Central Americans, Dominicans, and South Americans and other/multi), and body mass index (BMI) in HCHS/SOL; age, sex, study site (two sites with low sample sizes were combined), race (White versus “Non-White”, which consists of Hispanic, Black and Chinese Americans), and BMI in MESA. Model 2 (i.e., lifestyle model) adjusted for demographic and lifestyle variables – alcohol use, cigarette use, total physical activity (MET-min/day), and diet (Alternative Healthy Eating Index 2010) in HCHS/SOL; alcohol use and cigarette use in MESA. Model 3 (i.e., lifestyle and comorbidity model) adjusted for demographic, lifestyle and comorbidity variables - indicators for diabetes, hypertension, fasting insulin, fasting glucose, HOMA-IR, HDL, LDL, total cholesterol, triglycerides, systolic blood pressure and diastolic blood pressure in HCHS/SOL; hypertension, fasting glucose, HDL, LDL, cholesterol, triglycerides, systolic blood pressure and diastolic blood pressure in MESA. All models used the same set of individuals with complete set of sleep and covariate measures.

### Association analysis between individual metabolites and OSA and AHI

We tested the association of each of 219 metabolites (both continuous and dichotomized metabolites) with moderate-severe OSA and AHI in the HCHS/SOL. Each metabolite was the exposure in either linear or logistic regression (depending on the outcome) for each model. We accounted for the HCHS/SOL study design (sampling and clustering) and obtained representative effect estimates using survey regression implemented in the R survey package (4.0) (23). We controlled the false discovery rate (FDR) using the Benjamini-Hochberg procedure (24) and determined significant associations as those with FDR p-value<0.05. In the replication analysis, we tested the associations of these metabolites with OSA in logistic regression and with AHI in linear regression in MESA in models 1-3. We computed one-sided p-values guided by the estimated direction of associations in the HCHS/SOL (25), and determined replication if the one-sided p-value was <0.05.

### LASSO regression for constructing metabolite indices

We applied a LASSO logistic regression with moderate to severe OSA versus no or mild OSA (for brevity “OSA versus no OSA”), and linear regression with AHI – log transformed as log(AHI+1), adjusted for the covariates in Model 1 in HCHS/SOL. We included 209 continuous metabolites (not including the 10 dichotomized metabolites). We selected the LASSO tuning parameter by minimizing the misclassification error for OSA (assuming probability of 0.5 is the cutoff for “predicting” OSA), and the prediction error for AHI, in a 10-fold cross-validation. Metabolite indices were calculated as a weighted sum of the (normalized) metabolite serum concentrations, with weights being the metabolite coefficients from the LASSO regression.

To validate the metabolite index association with OSA/AHI, we constructed the indices in MESA using the weights from the LASSO regression conducted in HCHS/SOL, then assessed their associations with the corresponding sleep traits (i.e., OSA, AHI) in model 1-3. In the secondary analyses we assessed potential sex differences via: (1) sex-stratified association analysis for sex-specific metabolite indices (constructed based on sex-stratified LASSO), as well as (2) sex-stratified association analysis for metabolite indices constructed based on combined sexes. We also assessed the associations between metabolite indices quartiles and the corresponding sleep traits.

All analyses were done in R 3.6.3. The glmnet package (3.0)(26) in R was used for the LASSO logistic regression.

## Results

### Participant characteristics

**Table 1** characterizes the HCHS/SOL analytic sample and target population. The HCHS/SOL cohort included 3,507 participants, with a mean age of 41.72 years (SD =15.4), of whom 50.7% were female and 10.2% were classified with moderate or severe OSA (AHI ≥15). Participants with OSA were more likely to be male, had a higher BMI, and were less likely to be never smokers compared to those without OSA. Individuals with OSA were also more likely to have co-morbidities: 60% had hypertension and 34.3% had diabetes, compared to 27.8% hypertension and 16.9% diabetes in those without OSA. **S2 Table** characterizes the 475 MESA participants with metabolomics, sleep, and required measured covariates from the validation dataset. MESA participants were older (mean 68.45 years, SD=9.33), with a higher proportion of females (56.2%). Reflecting their older age, more MESA participants had moderate or severe OSA (46.7%) compared to HCHS/SOL.

**Table 1.**
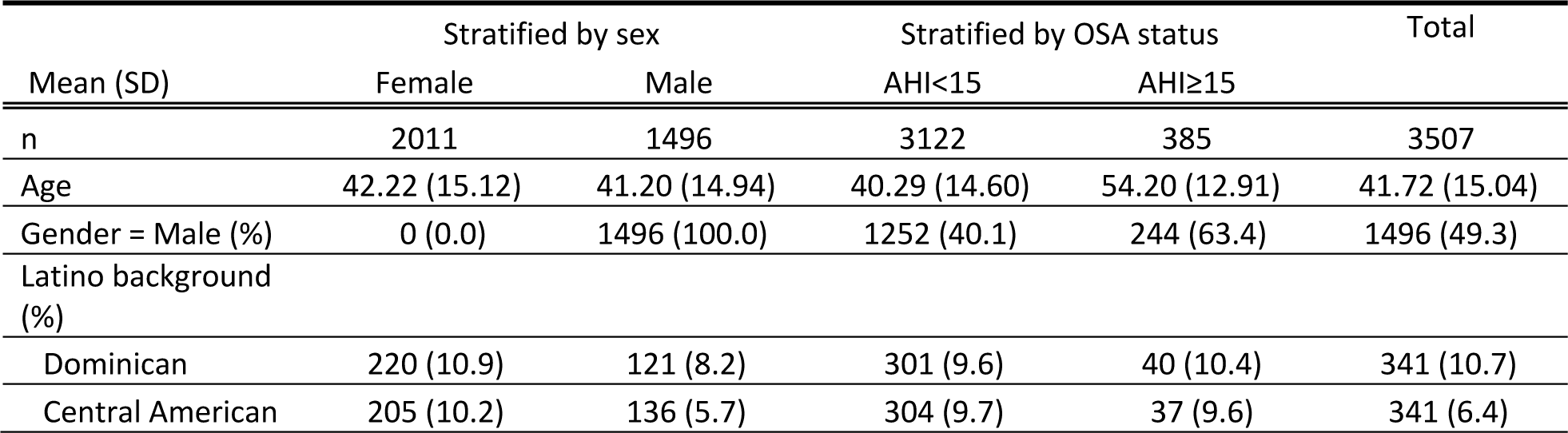
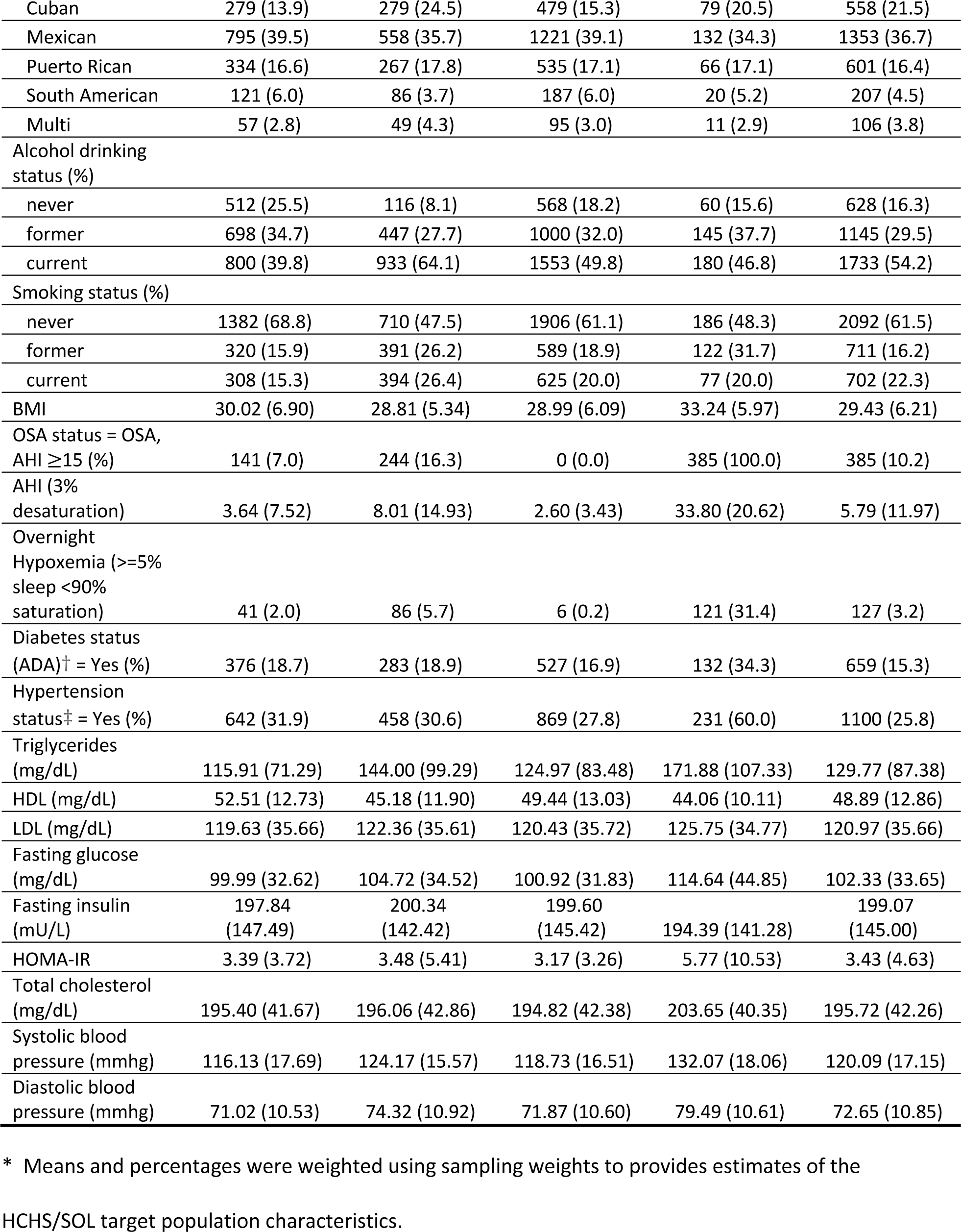
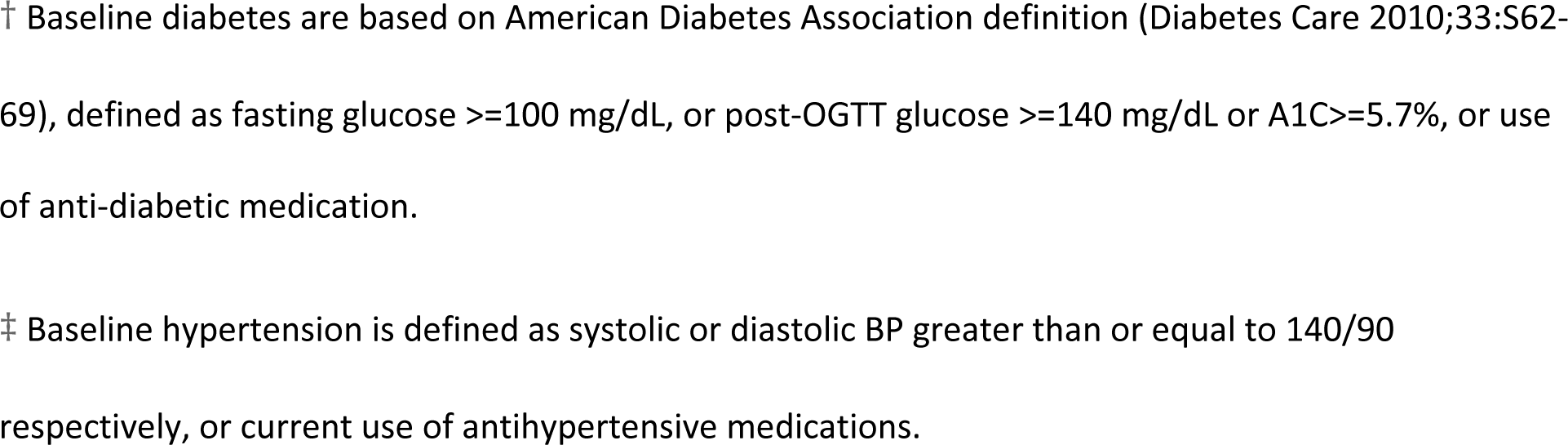
Characteristics of Hispanics/Latinos represented by the HCHS/SOL study population.

### Metabolite associations with OSA and AHI

**Table 2** shows the odds ratios corresponding to 7 metabolites associated (FDR *P* < 0.05) with OSA in HCHS/SOL adjusted for age, sex, BMI, study site/center, race/ethnicity). **Figure 2A** **and S3 Tables** show the lifestyle adjusted model and comorbidity adjusted model results. Among the 7 mapped metabolites in MESA, 4 metabolite associations had one sided p-values < 0.05 – glutamate, phenylalanine, linoleoyl-linoleoyl-glycerol (18:2/18:2), and oleoyl-linoeleoyl-glycerol (18:1/18:2), all of which were associated with increased risks for OSA (**Figure 2B**). These associations also had FDR p-value<0.05 in MESA. No metabolite was associated with AHI after multiple testing correction in the HCHS/SOL (**Supplemental Figure 2**). Also, no metabolite associations were detected at the FDR< 0.05 level in minimally adjusted sex-stratified analyses in HCHS/SOL.

**Figure 2.**
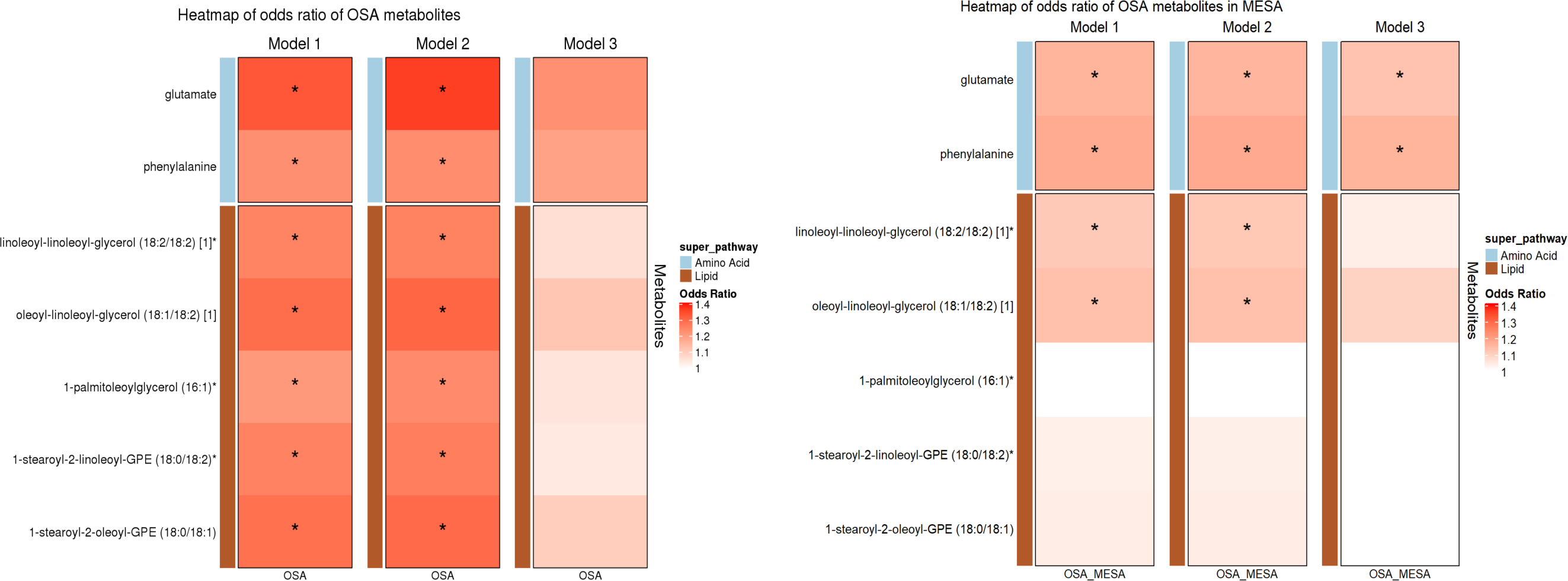
Heatmap showing estimated odds ratios of metabolites with significant FDR-adjusted p-value for OSA in HCHS/SOL and in MESA. * indicates FDR *p*<0.05. In HCHS/SOL: Model 1 adjusted for age, gender, center, background, and bmi. Model 2 adjusted for age, gender, center, background, bmi, alcohol use, smoking status, physical activity and diet (AHEI 2010). Model 3 adjusted for age, gender, center, background, bmi, alcohol use, smoking status, physical activity, diet, T2DM, hypertension, fasting glucose, fasting insulin, HOMA_IR, HDL, LDL, total cholesterol, triglycerides, systolic blood pressure and diastolic blood pressure. In MESA: Model 1 adjusted for age, gender, BMI, study site (site WFU and UCLA are combined due to low cell count), and race. Model 2 adjusted for age, gender, BMI, study site, race, alcohol use and smoking status. Model 3 adjusted for age, gender, BMI, study site, race, alcohol use, smoking status, hypertension indicator, fasting glucose, HDL, LDL, cholesterol, triglycerides, systolic blood pressure and diastolic blood pressure.

**Table 2.** Single metabolite associations with FDR-adjusted p-value <0.05 in the discovery step.

| Biochemical | Super-pathway | Sub-pathway | HMDB ID | HCHS/SOL |  |  | MESA |  |  |  |  |
| --- | --- | --- | --- | --- | --- | --- | --- | --- | --- | --- | --- |
|  |  |  |  | OR | Unadj-p<sup>†</sup> | FDR p | Platform | OR | Unadj-p<sup>†</sup> | 1-sided p | 1-sided FDR p |
| glutamate | Amino Acid | Glutamate Metabolism | HMDB00148 | 1.33 | 1.36E-04 | **0.014** | Amide-neg | 1.16 | 1.11E-02 | **0.006** | **0.019** |
| phenylalanine | Amino Acid | Phenylalanine Metabolism | HMDB00159 | 1.23 | 1.63E-03 | **0.049** | HIL-pos | 1.17 | 3.55E-03 | **0.002** | **0.014** |
| linoleoyl-linoleoyl-glycerol (18:2/18:2) [1]* | Lipid | Diacylglycerol | HMDB07248 | 1.25 | 8.27E-04 | **0.040** | HIL-pos | 1.11 | 3.47E-02 | **0.017** | **0.030** |
| oleoyl-linoleoyl-glycerol (18:1/18:2) [1] | Lipid | Diacylglycerol | HMDB07219 | 1.29 | 8.90E-05 | **0.014** | HIL-pos | 1.13 | 1.51E-02 | **0.008** | **0.019** |
| 1-palmitoleoylglycerol (16:1)* | Lipid | Monoacylglycerol | HMDB11565 | 1.21 | 1.15E-03 | **0.040** | C8-pos | 0.92 | 9.96E-02 | 0.950 | 0.950 |
| 1-stearoyl-2-linoleoyl-GPE (18:0/18:2)* | Lipid | Phosphatidylethanolamine (PE) | HMDB08994 | 1.25 | 9.89E-04 | **0.040** | HIL-pos | 1.03 | 5.37E-01 | 0.269 | 0.314 |
| 1-stearoyl-2-oleoyl-GPE (18:0/18:1) | Lipid | Phosphatidylethanolamine (PE) | HMDB08993 | 1.28 | 2.77E-04 | **0.019** | C8-pos | 1.04 | 4.49E-01 | 0.225 | 0.314 |

\* Metabolite with \* indicates they were identified based on accurate mass data, retention time and mass spectrometry but not reference
standards. Therefore, the verification is not as robust as metabolites without \*.
† All results were adjusted for age, gender, BMI, study center, and Hispanic/Latino background in HCHS/SOL, and adjusted for age, sex, BMI,
study site (site WFU and UCLA are combined due to low cell count), and race in MESA. Unadj-*p* is the abbreviations of raw *p* before the FDR
correction. 1-sided p-values reflect the required of the same direction of associations in the discovery and validation datasets for concluding
replication.

### LASSO regression for joint selection and estimation of metabolite associations with OSA and AHI in HCHS/SOL

We used a LASSO regression to select a set of metabolites that jointly associated with sleep apnea traits in the HCHS/SOL. Among the 14 metabolites identified for OSA by LASSO (**Figure 3**), there were one carbohydrate, one peptide, three amino acids, three lipids, three nucleotides, and three cofactors and vitamins, among which biliverdin and serine were unique to the OSA metabolite index while the rest were shared between the OSA and AHI metabolite indices. 41 metabolites were identified for AHI (**Supplemental Figure 3**), among which 29 metabolites were unique to AHI metabolite index. Metabolites identified by sex-specific LASSO are provided in the supplemental materials (**Supplemental Figures 4-7)**.

**Figure 3:**
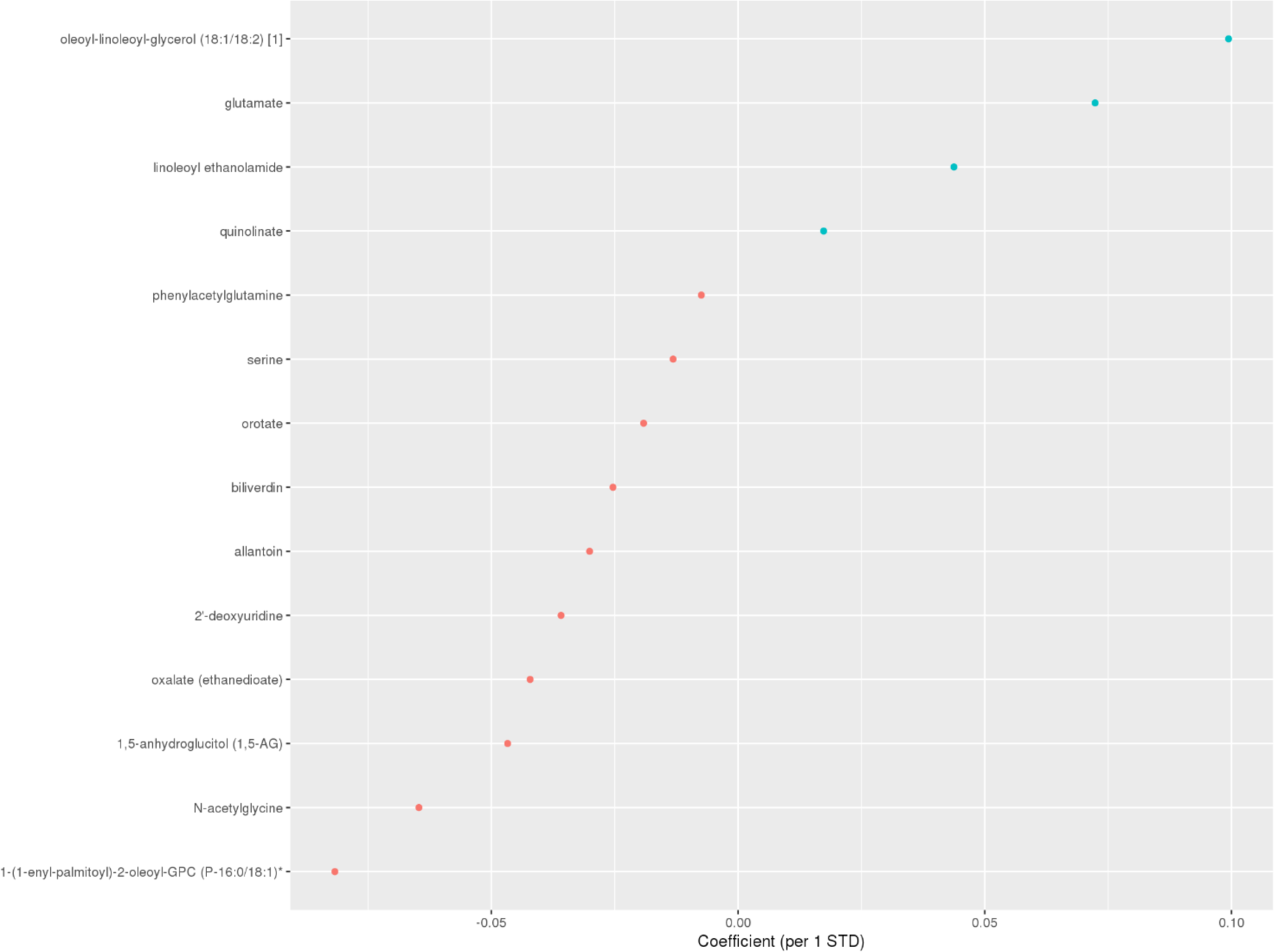
Coefficients for metabolites selected by LASSO (OSA model) in HCHS/SOL. Blue: coefficient<0; Red: coefficient>0. Definition of abbreviations: OSA = moderate to severe obstructive sleep apnea (AHI≥15), LASSO = least absolute shrinkage and selection operator.

### Metabolite indices associations with OSA and AHI in HCHS/SOL and in independent validation in MESA

We constructed OSA and AHI metabolite indices in both HCHS/SOL and MESA based on the weights from the LASSO regressions conducted in HCHS/SOL. **Table 3** provides overall and sex- stratified results. As expected by construction, the metabolite indices were associated with their phenotypes in HCHS/SOL (OSA metabolite index OR: 1.50; 95% CI 1.30-1.74; *P* < 0.001; AHI metabolite index beta =2.12 per 1 SD of the index, 95% CI 1.43-2.81; *P* < 0.001) after adjustment for demographic covariates (i.e., age, sex, BMI, study center, and Hispanic/Latino background); the associations persisted after additional adjustment of lifestyle (i.e., alcohol usage, smoking status, physical activity and diet) and comorbidity (i.e., diabetes, hypertension, fasting glucose, fasting insulin, HOMA-IR, HDL, LDL, total cholesterol, triglycerides, systolic blood pressure, diastolic blood pressure) covariates (OSA metabolite index OR: 1.50; 95% CI 1.21-1.85; *P* < 0.001; AHI metabolite index beta =1.73 per 1 SD of the index, 95% CI 0.96-2.50; *P*<0.001). In the validation dataset, the OSA metabolite index was also associated with an increased odds ratio for OSA (OR: 1.41; 95% CI 1.13-1.77; *P* = 0.003) (**Table 3**) when adjusted for demographic (i.e., age, sex, BMI, study site, and race) covariates and remained similar when adjusted for lifestyle (i.e., alcohol usage and smoking status) and comorbidity (i.e., hypertension, fasting glucose, HDL, LDL, cholesterol, triglycerides, systolic blood pressure and diastolic blood pressure) covariates (OR: 1.55; 95% CI 1.10-2.20; *P*= 0.013). When compared with the lowest quartile of the OSA metabolite index, the top quartile showed more than two-fold increase in risk for OSA [OR: 2.53; 95% CI (1.37-4.70); *P* = 0.003] in the primary model and remained significant when adjusted for lifestyle and comorbidity covariates [OR: 2.63; 95% CI (1.14-6.14); *P* = 0.024] (see **Figure 3** **and Supplemental Figure 3**). AHI metabolite index associations had higher p-values in MESA, compared to OSA metabolite index associations (**Table 3**). AHI metabolite index was only replicated in women, adjusted for demographic, lifestyle, and comorbidity covariates in MESA. Notably, both OSA metabolite index and AHI metabolite index associations with their phenotypes were stronger when evaluated in women compared to the overall sample, in both HCHS/SOL and MESA.

**Table 3.**
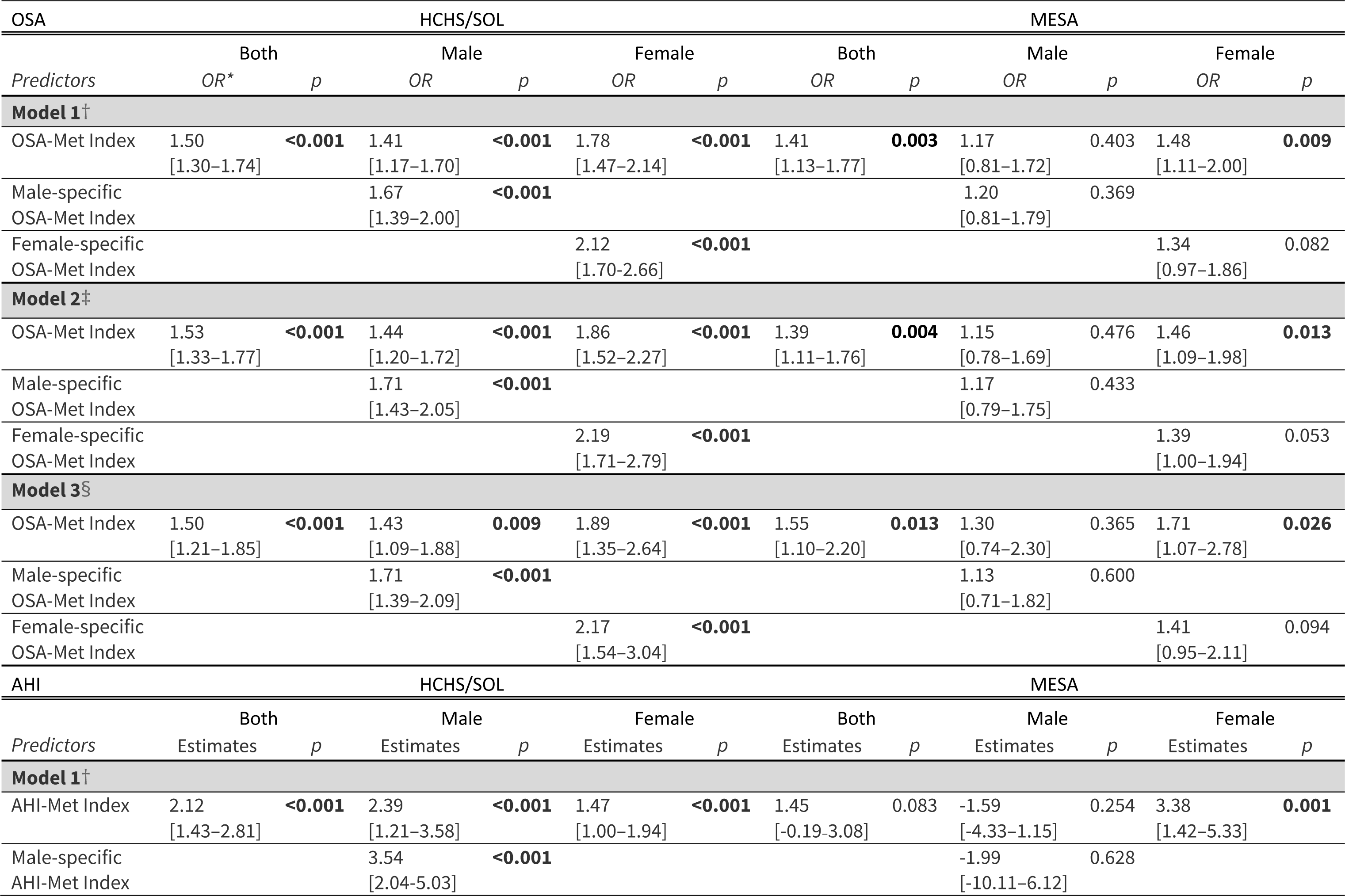
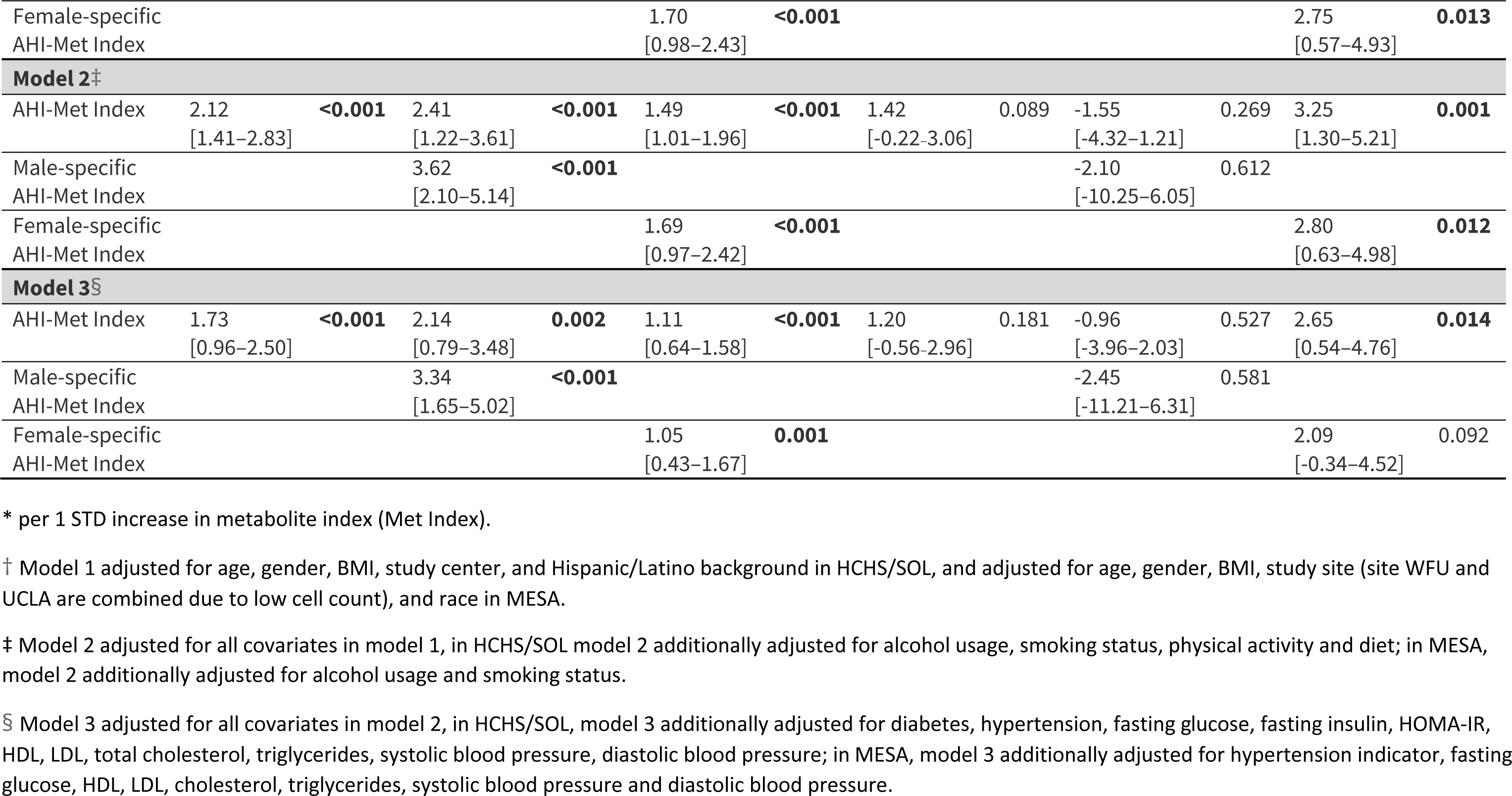
Estimated associations between OSA and AHI metabolite indices and their respective phenotypes, in HCHS/SOL and MESA.

Results from secondary analysis of a sex specific metabolite indices are also provided in **Table 3**. Only the female-specific AHI metabolite index replicated in MESA in model 1 and 2, but its association with AHI in women was weaker than that of the AHI metabolite index trained on the full HCHS/SOL sample, both in terms of p-value and of estimated effect size.

## Discussion

In this paper, we leveraged metabolomics data from two large, diverse community-based cohorts to derive the first metabolite index for moderate to severe OSA, as well as to identify individual metabolites associated with this disorder. We studied 219 metabolites and their associations with OSA and AHI in the HCHS/SOL using two methods: (1) analysis of individual metabolites, and (2) LASSO to identify a subset of metabolites that jointly predict OSA or AHI. Then, we studied the associations in an independent validation study, MESA. We used the results from LASSO to derive an OSA and AHI metabolite indices. In MESA, the OSA metabolite index was significantly associated with moderate to severe OSA; e.g., individuals in the highest quartile for OSA metabolite index had a more than 2-fold increased odds of moderate to severe OSA- both in the derivation sample and in an independent sample that varied by ancestry, age, and OSA prevalence, with findings that persisted after adjusting for multiple lifestyle and health covariates. In contrast, when modeling AHI as a continuous measure of sleep apnea, weaker associations were observed, except for the top quartiles among females. In the association analysis of individual metabolites, seven metabolites were associated with OSA in HCHS/SOL (FDR *p* < 0.05), of which four associations replicated in MESA.

We implemented two approaches to study the metabolomic correlates of sleep apnea phenotypes: LASSO and individual-metabolite regression analysis. These approaches serve different purposes: single metabolite regression highlights individual metabolites associated with sleep apnea phenotypes without adjustment to other metabolites, while LASSO estimates the combined effect of multiple metabolites. A metabolite identified as associated with sleep apnea phenotypes individually may not be selected by the LASSO analysis (e.g., if a different metabolite correlated to it was selected by LASSO). Thus, it is not surprising that two of the replicated metabolites identified in single metabolite analysis were not selected by LASSO. Similarly, a specific metabolite may be selected by LASSO, but not by individual metabolite analysis due to adjustment for multiple testing, which is not done in LASSO analysis. The OSA metabolite index, constructed based on the LASSO results, is a single index using multiple blood biomarkers which together reflect the biochemical differences in the blood of individuals with and without OSA. The OSA metabolite index also showed stronger association with OSA than any single metabolite, in both the discovery and validation study, consistent with the influence of multiple metabolites in OSA pathophysiology (see **Table 3, S3 Table**). While future work is needed to study whether the metabolite index can be used in the clinic for OSA screening or clinical management of OSA patients, it is clearly useful from a statistical and epidemiological standpoints, as analysis based on the metabolite index had evidently higher statistical power than analyses testing single metabolite associations. Therefore, it may also enable additional studies of the pathophysiology of OSA and its relationship with other cardiometabolic conditions.

Four metabolites were replicated in the single metabolite regression: glutamate, oleoyl-linoleoyl-glycerol (18:1/18:2) (DAG(36:3)), linoleoyl-linoleoyl-glycerol (18:2/18:2) (DAG(36:4)) and phenylalanine, among which glutamate and phenylalanine remained positively associated with OSA after adjusting for lifestyle and comorbidities in addition to basic demographics. Both metabolites have some previous evidence linking them to OSA or other sleep disorders, as well as cardiometabolic diseases, and suggest that elevations in glutamate and phenylalanine can be investigated as biomarkers for adverse outcomes in patients with OSA. High plasma glutamate has been associated with total and visceral adiposity, dyslipidemia and insulin resistance (27), as well as increased risks for incident cardiovascular disease (28), type 2 diabetes (29), and subclinical atherosclerosis (30), independent of established cardiovascular risk factors. The positive association between glutamate and OSA observed in our study may suggest a shared metabolomic profile for OSA and other cardiometabolic phenotypes. Previous studies in rats animals and human showed that more frequent sleep apneas led to increased level of glutamate in brain (31-33). Glutamate is the major excitatory neurotransmitter in the brain, and modulates brain energy metabolism and neuronal synaptic plasticity. Although the blood-brain barrier prevents plasma glutamate to freely permeate into the central nervous system (34), when glutamate increases in the brain, the brain-to-blood glutamate efflux also increases, as suggested by the correlation between the peripheral glutamate and the central nerve system glutamate levels (35). These findings support further research addressing the roles of peripheral and central glutamate in the pathophysiology of OSA.

Phenylalanine is an essential aromatic amino acid that plays a key role in the biosynthesis of other amino acids, including the neurotransmitters, dopamine, and norepinephrine. Studies have shown that plasma phenylalanine level can be elevated due to inflammation (36,37), and inflammation is a common finding in OSA (38). One mechanism for the increased levels of phenylalanine may be through chronic hypoxia, which has been reported to increase both systemic and cerebral delivery of phenylalanine (39). This is in line with our evidence: peripheral phenylalanine was elevated among individuals with moderate to severe OSA patients (**S3 Table**). A prior lab-based study that measured a few metabolites over the course of sleep reported that phenylalanine levels decreased less overnight among patients with OSA compared to controls (40). Phenylalanine levels were also reported to be elevated after sleep restriction (41). The downstream effects of phenylalanine have been studied more widely in other chronic conditions, with reports of associations with elevated pro-inflammatory cytokines, suppressed immunity and increased mortality among heart failure patients (42); more rapid telomere shortening consistent with accelerated aging (43). Recent studies reported elevations of phenylalanine associated with adverse COPD outcomes (44), which was postulated to reflect muscle breakdown and respiratory muscle insufficiency (45). Elevated plasma phenylalanine was shown to be a strong predictor for cardiovascular risk (46) and a biomarker, mediator and potentially therapeutic target for pulmonary hypertension (47). Further research on the association of OSA and phenylalanine may further identify the roles of hypoxia, inflammation, and muscle function in the pathophysiology of OSA and cardiometabolic conditions.

Our study has also shown that increased plasma levels of two diacylglycerols (DAGs): DAG(36:3) and DAG(36:4) were associated with moderate to severe OSA. Altered lipids metabolism is often observed among OSA patients (48-50); specifically, intermittent hypoxemia can stimulate lipolysis, increasing free fatty acid levels (51). Abnormalities in lipid metabolism may result in liver and skeletal muscle fat deposition, exacerbating OSA through inflammatory or muscle-related pathways (52). Therefore, the associations with these diacylglycerols may reflect mechanisms by which OSA related hypoxemia alters fatty acid metabolism. These associations, however, did not replicate in the validation study once adjusted for comorbidities, suggesting that the associations might be confounded by cardiometabolic conditions that often accompany OSA.

Estimated OSA associations, in both LASSO and single metabolite analysis, were generally stronger than AHI associations. Potential reasons are the variability of AHI and the potential non-linear metabolomic associations with AHI. Notably, non-linearity (i.e., a threshold effect) was previously shown for AHI association with hypoxemia and sympathetic nervous system activation burden (53).

Sex differences have been increasingly reported among OSA patients (17). Population-based studies have shown that the overall OSA prevalence is higher among men than women (54), while metabolic syndrome and cardiovascular conditions are more strongly associated with OSA among female patients compared to males (55,56). Indeed, when assessing the metabolite indices developed in both sexes and tested for their associations with OSA and AHI, we observed stronger associations among women than men in the MESA validation dataset (**Table 3**). However, compared to the metabolite indices developed in combined sex strata, sex-specific metabolite indices had weaker associations with the OSA/AHI in the validation data set (weaker effect size estimates and higher p-value), which may be the result of more misclassification when using a smaller dataset for discovery.

In addition to pointing to novel individual metabolites that play a role in OSA, our metabolite indices also showed moderately strong associations with OSA in an external sample, despite the marked differences in race/ethnicity, age, and OSA prevalence compared to the discovery sample. This supports the overall generalizability of the metabolite index across diverse populations. Nonetheless, the utility of a 2-fold increased risk of OSA among individuals in the highest metabolite index quartile in helping to screen or triage patients for more comprehensive testing will need to be formally evaluated, potentially combining metabolite data with other information, such as OSA-related symptoms, to improve screening.

A strength of our study is that our population-based sample is more than 10-fold larger than prior studies (57), includes a high proportion of ethnic/racial minorities who have been under-represented in research but are at increased for adverse health outcomes, and is more representative of samples in the general population who remain include large numbers of undiagnosed individuals. We used rigorous statistical methods, adjusted for a large number of lifestyle and health covariates, and were able to replicate the main findings despite large differences in our discovery and validation populations, which suggests relatively strong associations and generalizability of the metabolite associations with OSA.

There are several limitations in this study. The temporal relationship between the blood sample collection and sleep test was concurrent in HCHS/SOL and up to one year apart in MESA, allowing for cross-sectional associations, but limiting our ability to discern causal pathways.

Although over 1000 metabolites were quantified in both populations, less than 300 metabolites were matched between the two platforms (after quality control only 219 distinct metabolites were mapped). We limited our study to only the matched metabolites to allow for replication testing, which strengthens the results and conclusions. MESA metabolomic profiling was conducted using three complementary platforms measuring several broad classes of small molecules therefore multiple chemical compounds from MESA were mapped to the same metabolite in the HCHS/SOL. We chose a single feature to map to any HCHS/SOL metabolite based on a set of rules related presence of redundant ions, data missingness and skewness. In the future, other more optimal approaches may be proposed and studied. Some associations failed to replicate in MESA, potentially due to heterogeneity in different populations and low power in MESA, which had a small sample size. Finally, the definitions of AHI differed slightly in the two studies: while the 3% oxyhemoglobin desaturation criterion applied to hypopneas only in MESA, due to differences in the recording montage, a 3% desaturation criterion was applied for all respiratory events in HCHS/SOL.

In summary, we used two large datasets of population-based multi-ethnic cohort studies to study metabolomics associations with OSA. We developed an metabolite index that replicated across datasets, and had statistically significant association with OSA even after adjustment to cardiometabolic comorbidities. In future work we will study the possibility of developing an OSA screening tool based on this metabolite index. Four metabolites also replicated in an independent data set, of which one was previously implicated in OSA, and two were previously connected to sleep disorders. Collectively, our findings support the utility of metabolomic profiling to generate metabolite indices of sleep apnea in racially diverse populations, and to OSA’s pathophysiology.

## Data Availability

MESA and HCHS/SOL data are available through application to dbGaP according to
the study specific accessions. MESA phenotypes are available in: phs000209, and
HCHS/SOL phenotypes: phs000810. META metabolomics data will become available on dbGaP via the "NHLBI TOPMed: Multi-Ethnic Study of Atherosclerosis (MESA)" project (accession phs001416). HCHS/SOL metabolomics data are available via data use agreement with the HCHS/SOL Data Coordinating Center at the University of North Carolina at Chapel Hill, see collaborators website: https://sites.cscc.unc.edu/hchs/.

## Acknowledgement

The authors thank the staff and participants of HCHS/SOL and MESA for their important contributions. We gratefully acknowledge the studies and participants who provided biological samples and data for TOPMed.

## Data availability statement

MESA and HCHS/SOL data are available through application to dbGaP according to the study specific accessions. MESA phenotypes are available in: phs000209, and HCHS/SOL phenotypes: phs000810. HCHS/SOL metabolomics data are available via data use agreement with the HCHS/SOL Data Coordinating Center at the University of North Carolina at Chapel Hill, see collaborators website: https://sites.cscc.unc.edu/hchs/.

## Supporting information

**S1 Table. Mapping between MESA and HCHS/SOL Metabolomics Platform.** The mapping was carried out before any quality control steps (e.g. assessment of missingness, etc.). 294 compounds in MESA were mapped to 231 metabolites in HCHS/SOL. 60 redundant features in MESA were dropped based on the principles described in the methods section. After discarding 12 metabolites with high missingness(>=75%) in HCHS/SOL, 209 metabolites with low missingness (<25%) were imputed and 10 metabolites with medium missingness (25-75%) were dichotomized in the HCHS/SOL metabolomics platform. We applied the same QC steps to the corresponding metabolites in MESA, i.e., discarded, imputed, and dichotomized, the same metabolites in MESA as in HCHS/SOL.

**S2 Table. Characteristics of the analytic sample from the MESA study population.** * Baseline hypertension is defined as systolic blood pressure (SBP) > 130 mmHg, diastolic blood pressure (DBP) > 80 mmHg or any history of antihypertensive medication intake.

**S3 Table. Single metabolite associations with FDR-adjusted p-value <0.05 in HCHS/SOL and MESA.** Model 1 adjusted for age, gender, BMI, study center, and Hispanic/Latino background in HCHS/SOL, and adjusted for age, gender, BMI, study site (site WFU and UCLA are combined due to low cell count), and race in MESA. Model 2 adjusted for all covariates in model 1, in HCHS/SOL model 2 additionally adjusted for alcohol usage, smoking status, physical activity and diet; in MESA, model 2 additionally adjusted for alcohol usage and smoking status. Model 3 adjusted for all covariates in model 2, in HCHS/SOL, model 3 additionally adjusted for diabetes, hypertension, fasting glucose, fasting insulin, HOMA-IR, HDL, LDL, total cholesterol, triglycerides, systolic blood pressure, diastolic blood pressure; in MESA, model 3 additionally adjusted for hypertension indicator, fasting glucose, HDL, LDL, cholesterol, triglycerides, systolic blood pressure and diastolic blood pressure. Metabolite with * indicates they were identified based on accurate mass data, retention time and mass spectrometry but not reference standards.

Therefore, the verification is not as robust as metabolites without *.

**S4 Table. Estimated associations between OSA and AHI metabolite indices and their respective phenotypes in quartiles, in HCHS/SOL and MESA.** * per 1 STD increase in metabolite index. Model 1 adjusted for age, gender, BMI, study center, and Hispanic/Latino background in HCHS/SOL, and adjusted for age, gender, BMI, study site (site WFU and UCLA are combined due to low cell count), and race in MESA. Model 2 adjusted for all covariates in model 1, in HCHS/SOL model 2 additionally adjusted for alcohol usage, smoking status, physical activity and diet; in MESA, model 2 additionally adjusted for alcohol usage and smoking status. Model 3 adjusted for all covariates in model 2, in HCHS/SOL, model 3 additionally adjusted for diabetes, hypertension, fasting glucose, fasting insulin, HOMA-IR, HDL, LDL, total cholesterol, triglycerides, systolic blood pressure, diastolic blood pressure; in MESA, model 3 additionally adjusted for hypertension indicator, fasting glucose, HDL, LDL, cholesterol, triglycerides, systolic blood pressure and diastolic blood pressure.

