## Supplemental figures for "Development and Validation of a Metabolite Index for Obstructive Sleep Apnea across Race/Ethnicities"

Supplemental Figure 1: Flowchart of the study population selection and metabolomic data preprocessing.

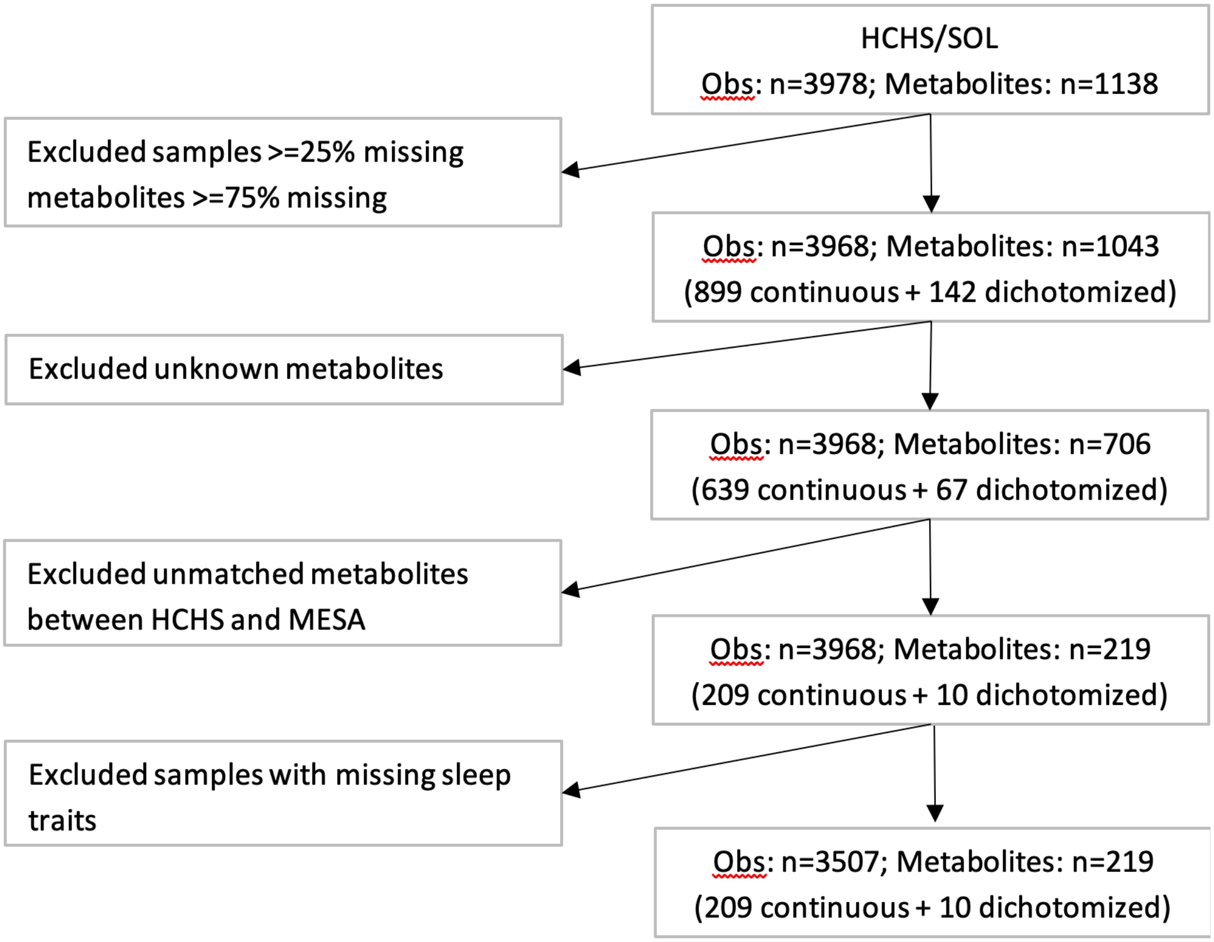

Definition of abbreviations: Obs=observations

### Supplemental Figure 2: Heatmap showing estimated coefficients of metabolites with significant FDR-adjusted p-value for AHI in HCHS/SOL.

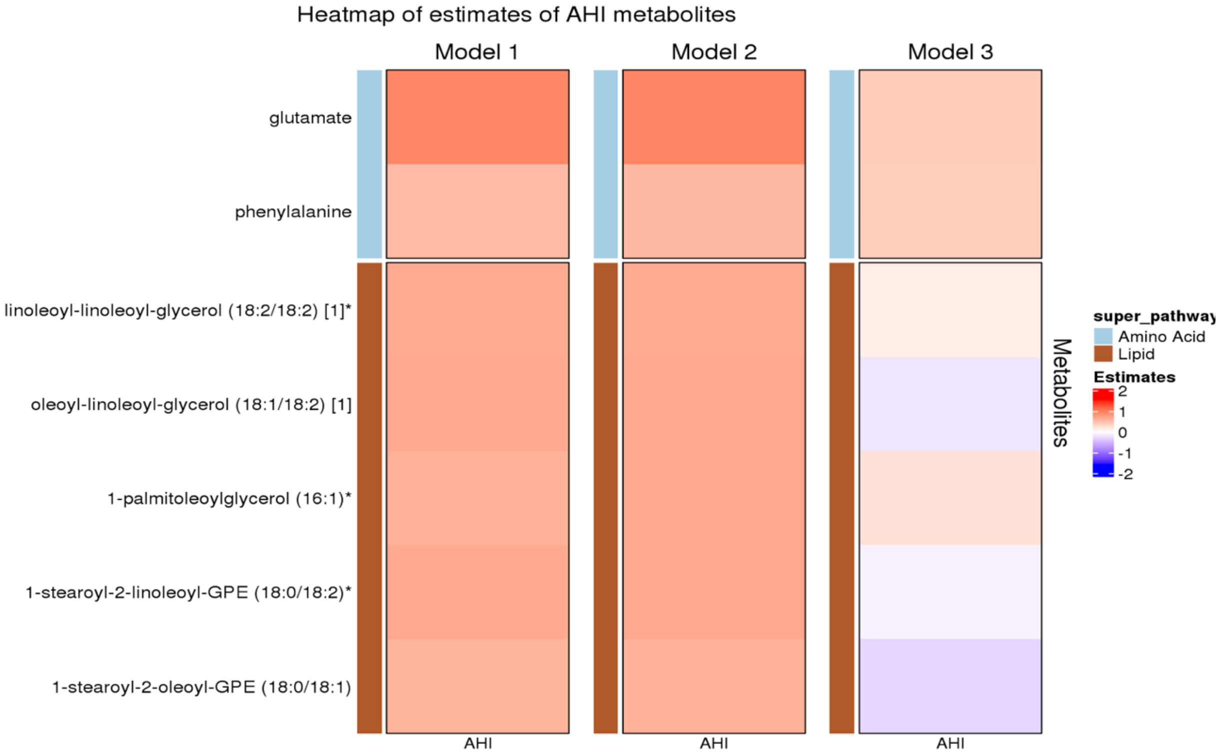

* indicates FDR p<0.05. Model 1 adjusted for age, gender, center, background, and bmi. Model 2 adjusted for age, gender, center, background, bmi, alcohol use, smoking status, physical activity and diet (AHEI 2010). Model 3 adjusted for age, gender, center, background, bmi, alcohol use, smoking status, physical activity, diet, T2DM, hypertension, fasting glucose, fasting insulin, HOMA_IR, HDL, LDL, total cholesterol, triglycerides, systolic blood pressure and diastolic blood pressure.

### Supplemental Figure 3: Coefficients for metabolites selected by LASSO (AHI model)

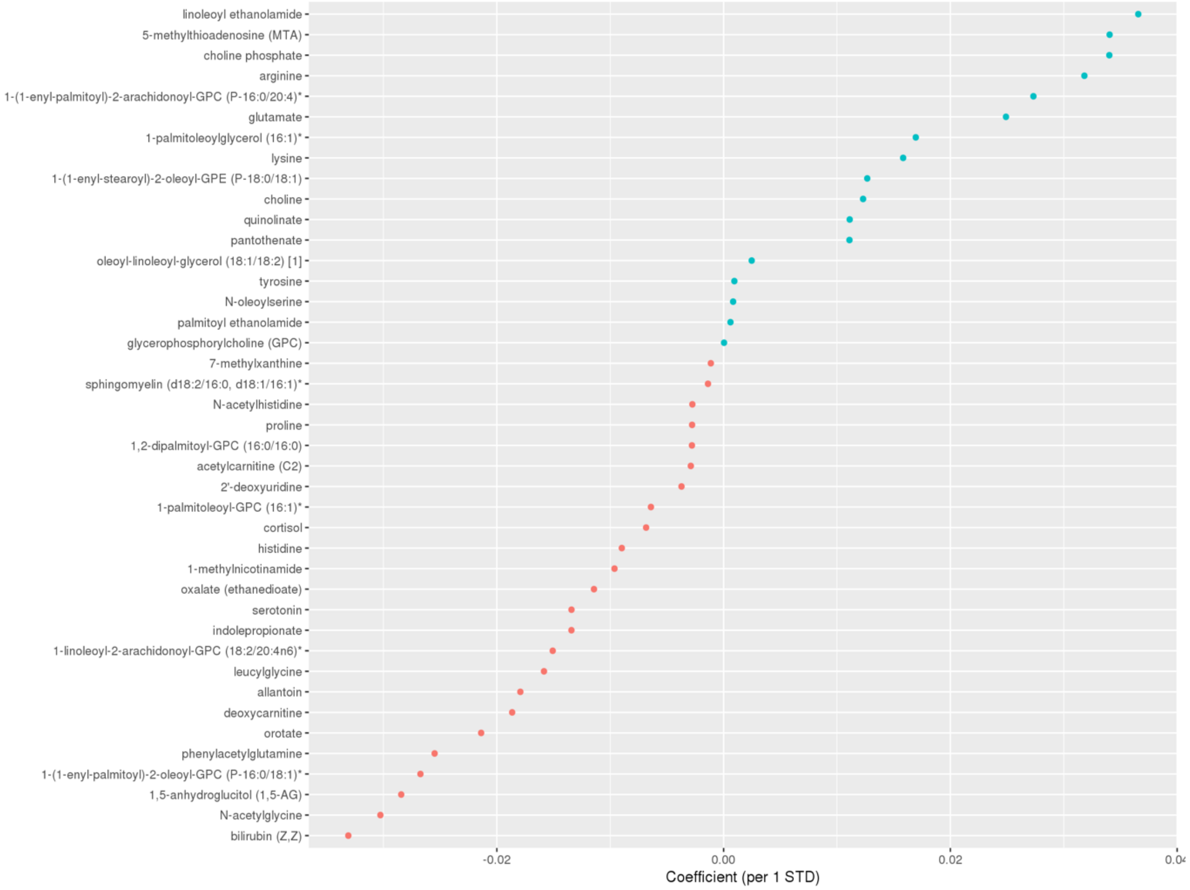

Blue: coefficient<0; Red: coefficient>0. Definition of abbreviations: OSA = moderate to severe obstructive sleep apnea (AHI≥15), LASSO = least absolute shrinkage and selection operator.

### Supplemental Figure 4: Coefficients for metabolites selected by LASSO among males only (OSA model)

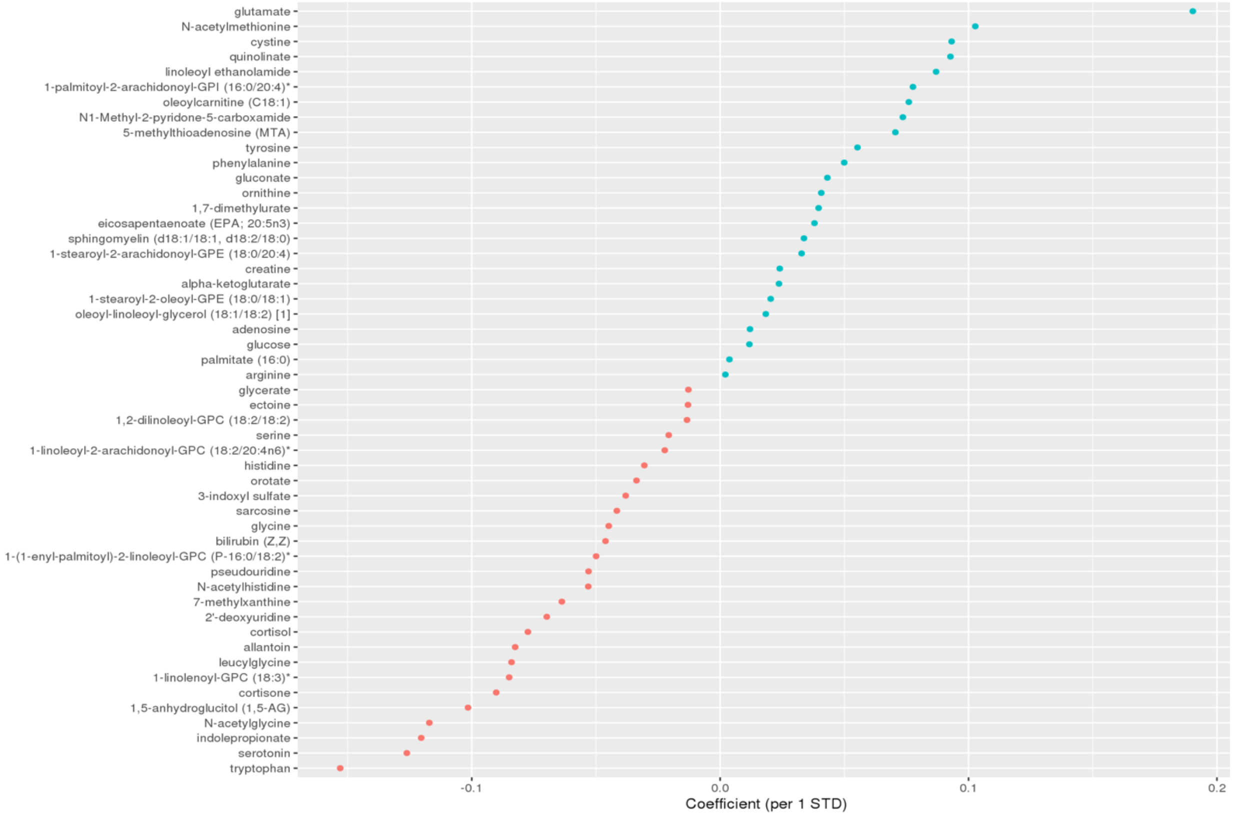

Blue: coefficient<0; Red: coefficient>0. Definition of abbreviations: OSA = moderate to severe obstructive sleep apnea (AHI≥15), LASSO = least absolute shrinkage and selection operator.

Supplemental Figure 5: Coefficients for metabolites selected by LASSO among females only (OSA model)

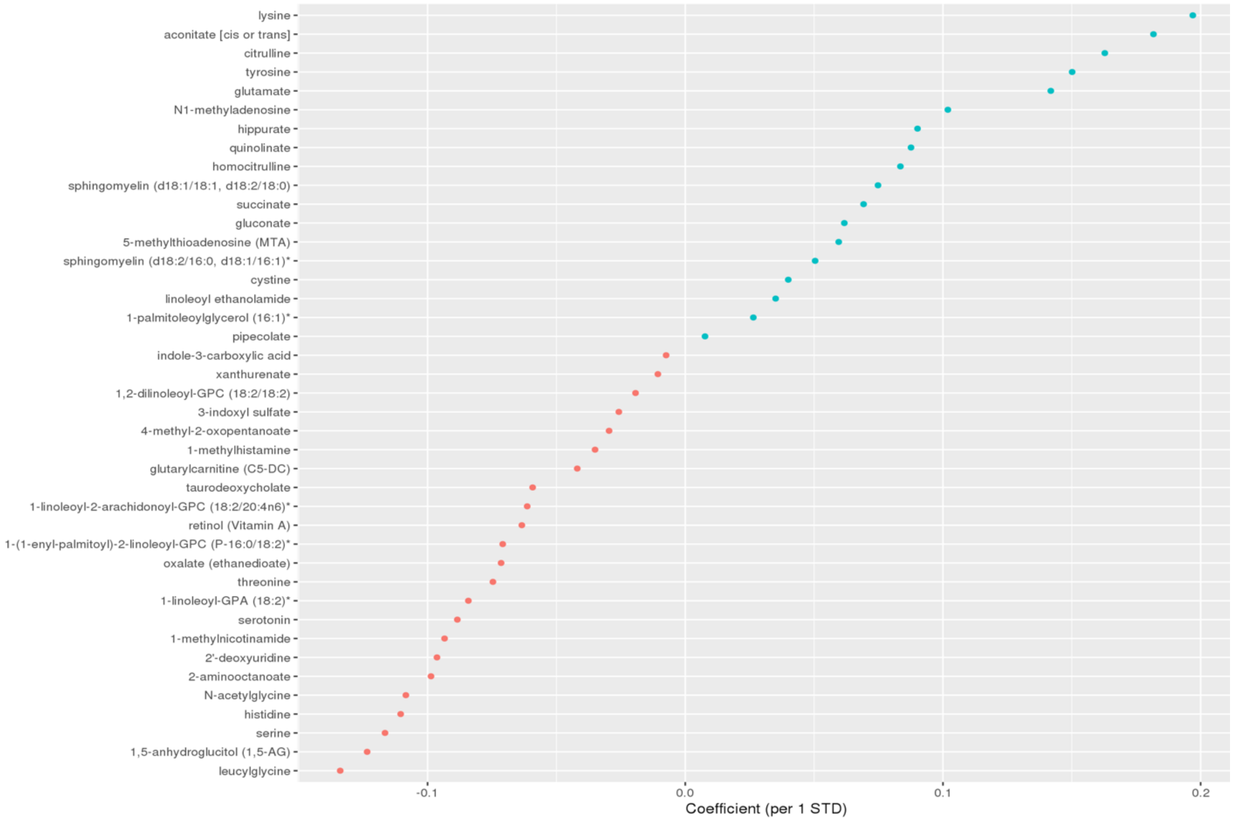

Blue: coefficient<0; Red: coefficient>0. Definition of abbreviations: OSA = moderate to severe obstructive sleep apnea (AHI≥15), LASSO = least absolute shrinkage and selection operator.

### Supplemental Figure 6: Coefficients for metabolites selected by LASSO among males only (AHI model)

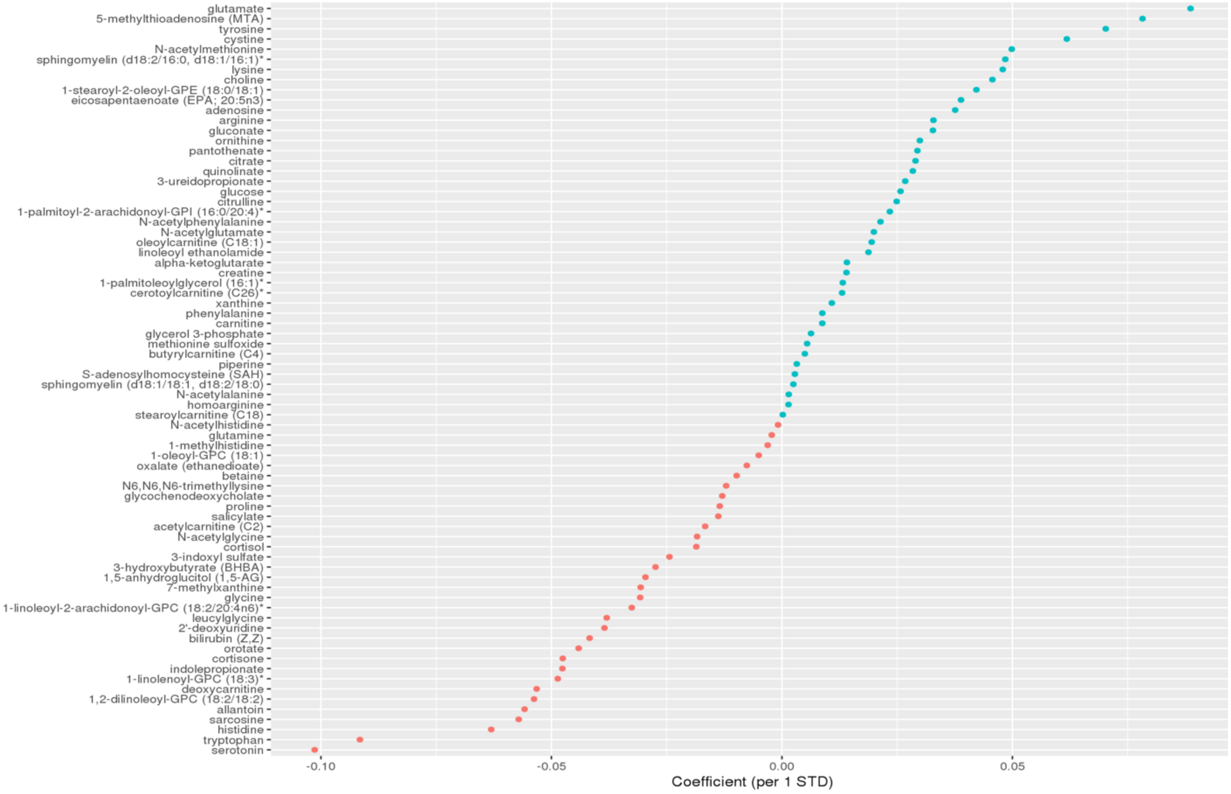

Blue: coefficient<0; Red: coefficient>0. Definition of abbreviations: OSA = moderate to severe obstructive sleep apnea (AHI≥15), LASSO = least absolute shrinkage and selection operator.

### Supplemental Figure 7: Coefficients for metabolites selected by LASSO among females only (AHI model)

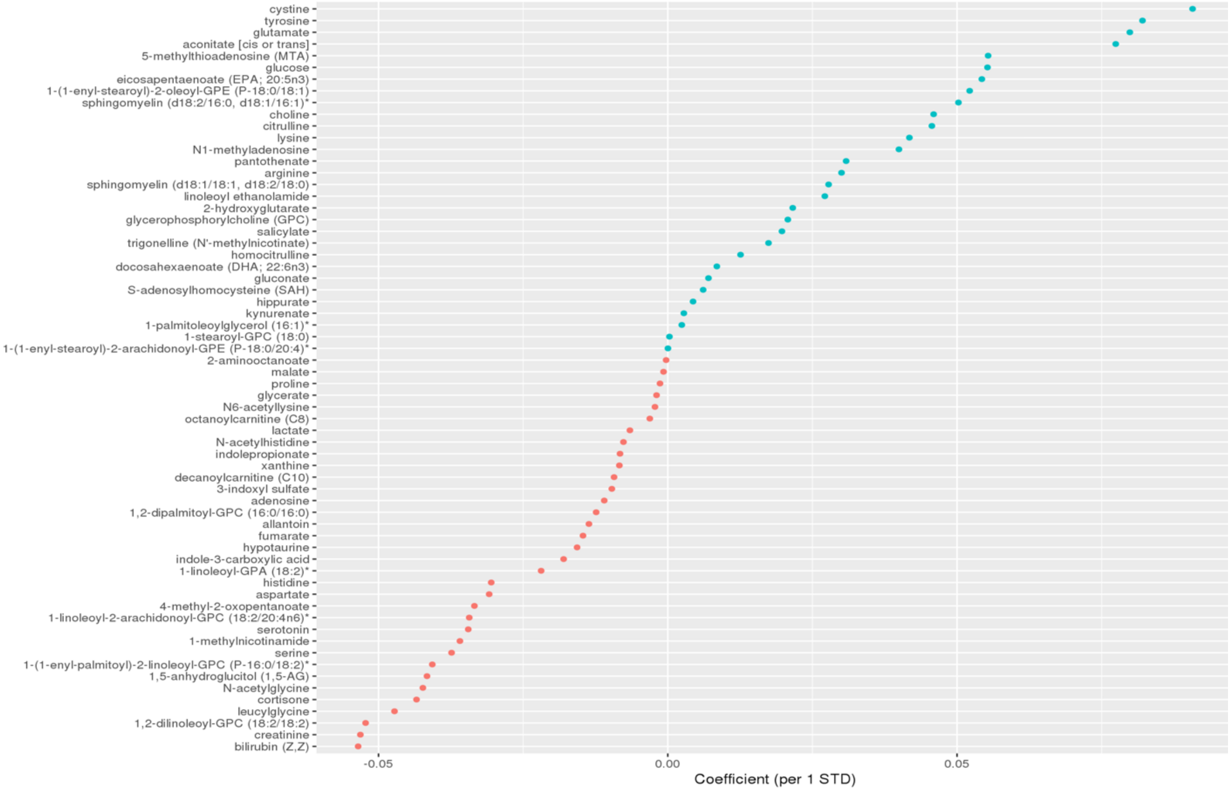

Blue: coefficient<0; Red: coefficient>0. Definition of abbreviations: OSA = moderate to severe obstructive sleep apnea (AHI≥15), LASSO = least absolute shrinkage and selection operator.

Supplemental Figure 8: percentage of moderate to severe OSA cases by OSA metabolite index quartiles in MESA.

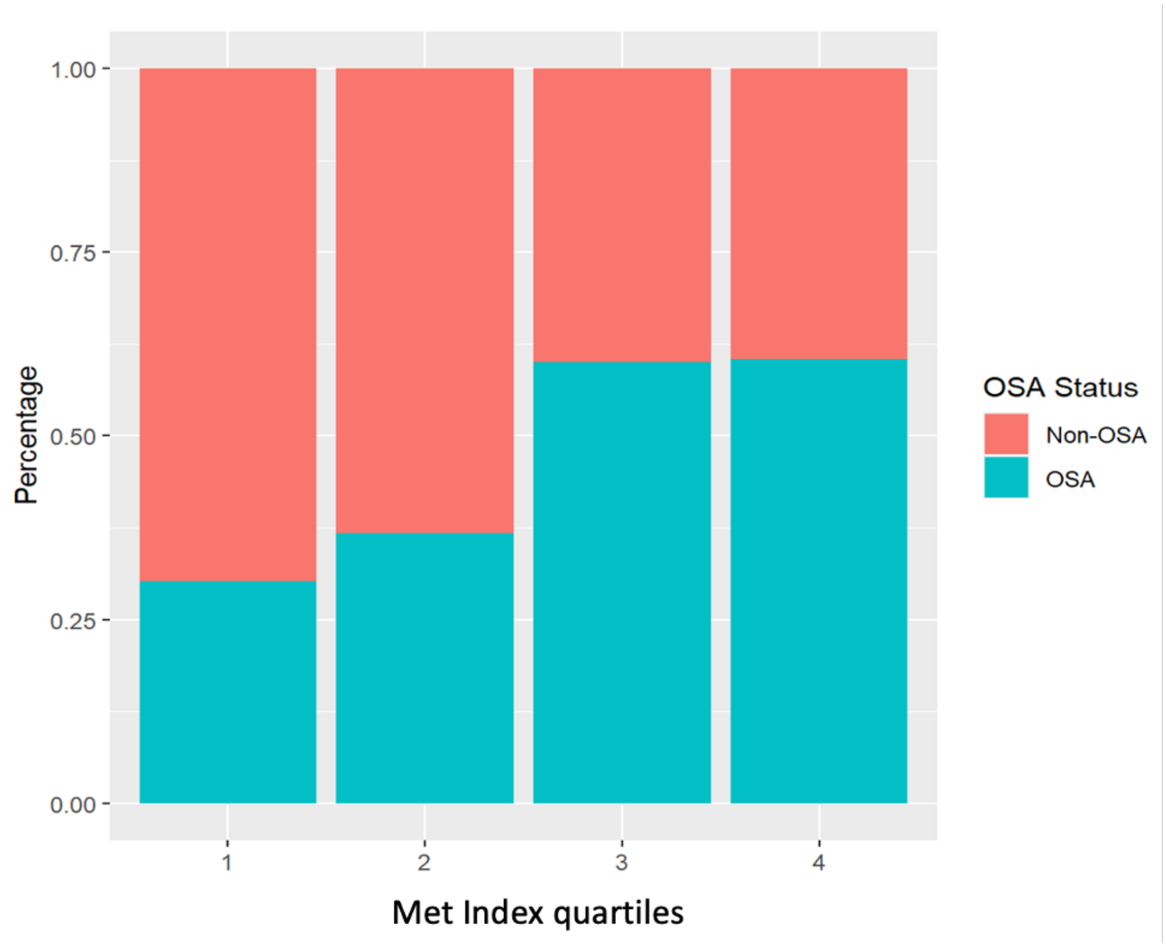
